# Proton Pump Inhibitors Prescribing Behaviors and Rationalisation Strategies among Healthcare Providers in the Southeast Asia

**DOI:** 10.1101/2025.10.15.25338052

**Authors:** Duc Trong Quach, Tien Manh Huynh, Augusto Jose G. Galang, Hang Viet Dao, Sakkarin Chirapongsathorn, Phuripong Kijdamrongthum, Rahela Ambaras Khan, Andrew Ming-Liang Ong, Wee Kooi Cheah, Kalwinder Singh Khaira, So Fie Tan, Jose Sollano, Ari Fahrial Syam, Tju Siang Chua, Somchai Leelakusolvong, Yeong Yeh Lee

**Author notes:** Corresponding author: **Professor Yeong Yeh Lee,** School of Medical Sciences, Universiti Sains Malaysia, Kota Bharu, Malaysia., **Professor Somchai Leelakusolvong**, Division of Gastroenterology, Department of Medicine, Faculty of Medicine Siriraj Hospital, Mahidol University, Bangkok, Thailand.

## Abstract

**Background:** Despite its widespread use, real-world evidence on proton pump inhibitor (PPI) prescribing and rationalisation practices in Southeast Asia (SEA) remains limited. This study aimed to investigate current prescribing behaviors and rationalisation strategies among SEA healthcare professionals.

**Methods:** The SEA PPI Rationalisation Working Group designed and disseminated an online survey across SEA via professional networks. The questionnaire included sections on demographics, PPI prescribing habits, and rationalisation strategies. Reliability testing of the questionnaire yielded a Cronbach’s alpha of 0.799. Content validity was confirmed by a panel of 10 gastroenterology experts, with perfect agreement on relevance and clarity (I-CVI, kappa, SCVI/Ave = 1.0). K-mode clustering was used to explore prescribing patterns. The main outcome was the identification of prescribing patterns and rationalisation strategies for long-term PPI use.

**Findings:** Among the 869 responses received, 763 valid entries were analysed (response rate: 87.8%), with 49% of the respondents being gastroenterologists. PPIs were most prescribed for gastroesophageal reflux disease (67.8%) and peptic ulcer disease (43.8%). Long-term PPI use was reported by 48.9% of the participants, while 59.2% reported reassessing indications at every follow-up visit. Rationalisation strategies included dose tapering (46.8%), on-demand use (45.9%), and step-down to antacids or alginates (44.8%). Cluster analysis identified two profiles: proactive prescribers (n=326, 42.7%) who followed clinical guidelines, applied individualised dosing, and routinely reassessed therapy, and conservative prescribers (n=437, 57.3%) who used PPIs more restrictively and rationalised less frequently.

**Interpretation:** Marked variability in PPI prescribing and rationalisation across SEA highlights the need for regional consensus to support evidence-based practice.

**Funding:** This study was supported by *Reckitt Benckiser (Singapore) Pte Ltd*., which provided funding for article processing charge

**Evidence before this study:** Although proton pump inhibitors (PPIs) are widely prescribed across Southeast Asia (SEA), there is limited real-world evidence examining the prescribing behaviors and rationalisation practices within the region. Previous research predominantly focused on Western countries and raised concerns regarding the over-prescription and prolonged use of PPIs. However, no studies have comprehensively addressed the regional variability in PPI prescribing or the strategies employed to rationalise their use in SEA.

**Added value of this study:** This study provides the first region-wide analysis of PPI prescribing practices and rationalisation strategies in SEA. By employing cluster analysis, the study identifies two distinct prescriber profiles: proactive prescribers, who follow clinical guidelines and individualise treatment, and conservative prescribers, who use PPIs more restrictively and apply rationalisation strategies less frequently. The study offers critical insights into the current prescribing landscape in SEA, highlighting the need for evidence-based regional guidelines to optimise PPI use.

**Implications of all the available evidence:** The findings suggest that significant variability in PPI prescribing exists across SEA, underscoring the necessity for region-specific guidelines. Policymakers should use these insights to inform evidence-based clinical guidelines for PPI use and rationalisation. Additionally, future research should explore the effectiveness of rationalisation strategies and identify barriers to their implementation within diverse healthcare systems in SEA.

## INTRODUCTION

Proton pump inhibitors (PPIs) are the mainstay in the treatment of acid-related gastrointestinal disorders. The clinical efficacy of these drugs has led to widespread global prescription. Nevertheless, the inappropriate and prolonged use of PPIs has become a growing concern. Several studies have reported significantly inappropriate prescribing rates in primary care and hospital inpatients.^1,2^ In Southeast Asia (SEA), this issue is particularly pronounced, with several studies in the region reporting inappropriate PPI prescribing rates ranging from 40% to 60%.^3–5^ Long-term PPI use has raised concerns about its potential association with a spectrum of adverse clinical outcomes, including *Clostridioides difficile* infection, small intestinal bacterial overgrowth, chronic kidney disease, bone fractures, and micronutrient deficiencies, although the current level of evidence remains low.^6^ However, acid-related gastrointestinal disorders in Asian populations are somewhat different from those in their Western counterparts. Notably, although the prevalence of gastroesophageal reflux disease (GERD) is increasing in the region, cases of erosive reflux disease are generally mild, and endoscopic complications remain uncommon.^7^ These scenarios typically do not necessitate long-term PPI maintenance therapy. These findings suggest that PPI prescribing patterns in clinical practice across Southeast Asia may differ substantially from those observed in Western healthcare settings. However, despite these differences, regional data remain sparse and fragmented, and there is still no unified consensus on deprescribing protocols tailored to the region. In this context, the present study aimed to evaluate current PPI prescribing behaviours and rationalisation strategies among healthcare providers across Southeast Asia. The findings are intended to support the development of evidence-based, regionally tailored recommendations to optimise the appropriate use of PPIs within the region’s varied and dynamically changing healthcare environment.

## METHODS

### Survey development and validation

The questionnaire was developed by the SEA PPI Rationalisation Working Group. The content development was based on three core expert insights from the group and review of existing literatures using pre-defined search criteria. Initial iterations of the newly developed questionnaire were then distributed to three other experts for comments and revised accordingly. The final English language-based questionnaire comprises 13 structured items across three sections: (1) demographic and professional background; (2) PPI prescribing habits and perceptions; and (3) PPI rationalisation strategies (Supplement 1). Content validity was evaluated by a panel of 10 regional experts, with each item assessed via the content validity index (CVI).^8^ A pilot test involving 30 healthcare professionals was conducted to evaluate clarity, item sequence, and overall comprehensibility.

### Online study design and participants

The survey was decided by the working group to distribute online and using cross-sectional design. It was distributed using Survey Monkey (website: https://www.surveymonkey.com/) between May and August 2025 in six Southeast Asian countries: Indonesia, Malaysia, the Philippines, Singapore, Thailand, and Vietnam. The participants were healthcare professionals who were members of national societies in gastroenterology, internal medicine, geriatrics, or pharmacy. The eligibility criteria were as follows: (1) active membership in at least one of these professional societies and (2) provision of informed consent by agreeing to participate at the beginning of the survey. The respondents were recruited through the official communication channels of the respective societies. In addition, targeted email invitations were sent by core members of the SEA PPI Rationalisation Working Group, who many also served as executive members of the participating national societies. This recruitment strategy aimed to ensure diverse professional representations and enhance response validity across countries. Only fully completed questionnaires were included in the final dataset, and duplicate submissions identified through identical IP addresses were excluded.

### Statistical analysis

Descriptive statistics were used to summarise the demographic and survey response data including categorical variables (frequencies and percentages) and continuous variables (means with standard deviations or medians with interquartile ranges). Group comparisons were performed across two key subgroups: (i) gastroenterology (GI) versus non-GI healthcare professionals (all other specialties) and (ii) high-level healthcare experience (HE; defined as ≥10 years in clinical practice) versus low-level experience (LE; <10 years). For comparisons involving categorical variables, either Pearson’s chi-square test or Fisher’s exact test was applied, as deemed appropriate.

K-mode clustering, an unsupervised machine learning algorithm suitable for categorical data, was used to explore latent behavioural patterns.^9^ The respondents were grouped based on similarities across multiple survey domains, including indications for long-term PPI use, deprescribing strategies, and adjunctive therapies. The optimal number of clusters was determined using the elbow method based on within-cluster dissimilarity. To identify predictors of proactive prescribing behavior, univariate logistic regression analyses were conducted to estimate crude odds ratios (ORs) with corresponding 95% confidence intervals (CIs). Variables with p values <0.05 or those considered clinically meaningful were subsequently included in a multivariable logistic regression model to estimate adjusted ORs and 95% CIs. All analyses were performed via R version 4.4.0 (R Foundation for Statistical Computing, Vienna, Austria). Statistical significance was defined as a two-sided p-value <0.05.

### Ethical considerations

The study obtained approval from the Institutional Review Board of the University of Medicine and Pharmacy at Ho Chi Minh City, Vietnam (approval number: 2579/DHYD-HDDD). It was conducted in accordance with the ethical principles outlined in the Declaration of Helsinki.

## RESULTS

### Survey validation

The newly developed survey achieved a scale-CVI/average of 1.0, indicating excellent content validity. From pilot testing, the final questionnaire demonstrated good internal consistency, with a Cronbach’s alpha of 0.799.

### Cross-sectional online survey

A total of 869 responses were collected via the online survey platform, of which 763 met the eligibility criteria and were included in the final analysis (Figure 1). The baseline characteristics of participants are presented in Table 1. Nearly half of the participants were GIs and half with more than 10 years of clinical experience (49.4% and 48.3%, respectively). Majority of participants were hospital-based, with 42.6% working in government hospitals and 46.9% working in private hospitals.

**Figure 1.**
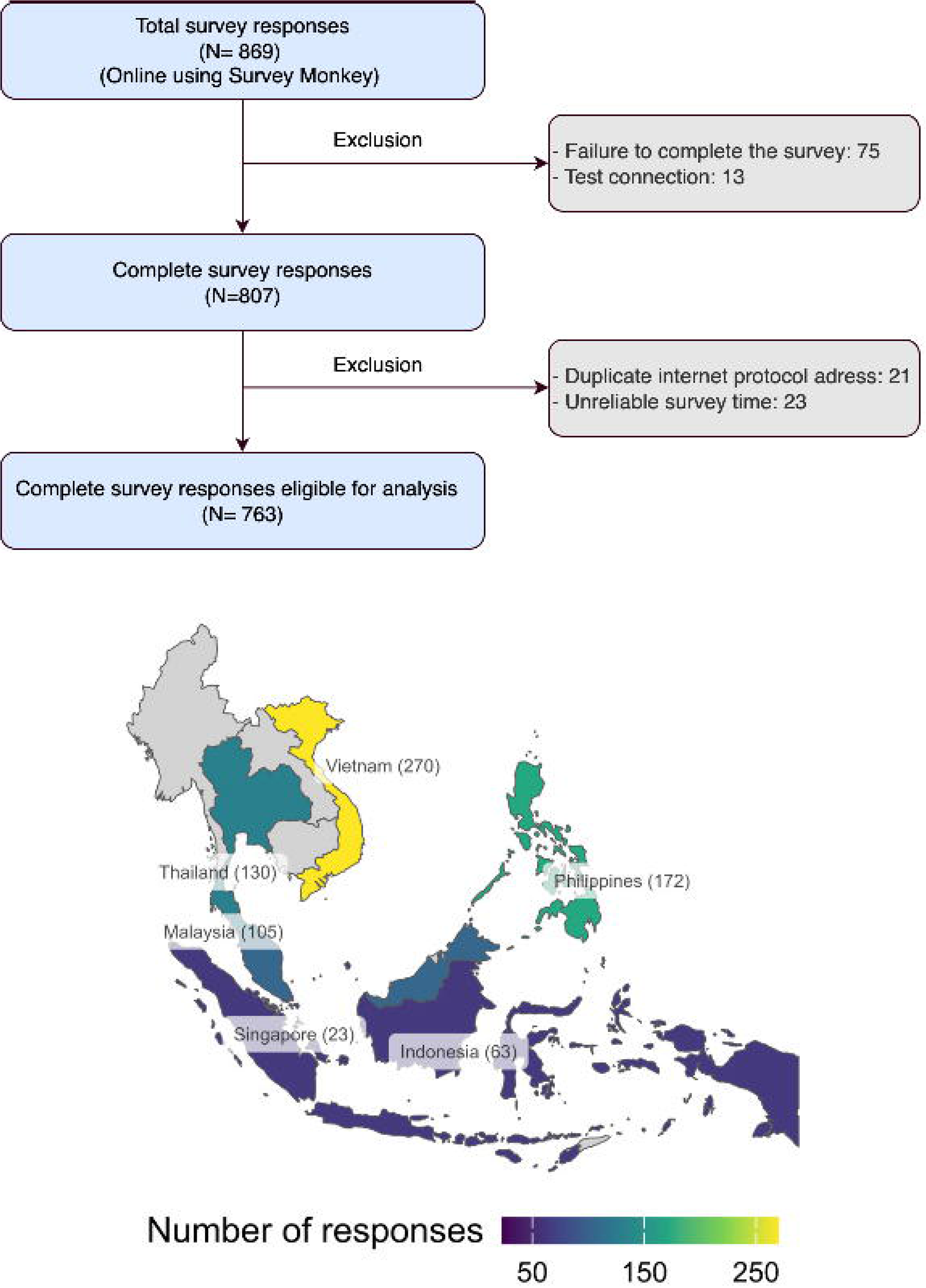
Study flow and geographic distribution of survey respondents.

**Table 1.** Baseline characteristics of survey respondents (N=763)

| Characteristic | n (%) |
| --- | --- |
| Specialty |  |
| Gastroenterology | 377(49.4) |
| General practitioner/family physician | 145 (19) |
| Geriatric medicine | 10 (1.3) |
| Internal medicine | 123 (16.1) |
| Obstetrics and gynaecology | 2 (0.3) |
| Otolaryngology | 68(8.9) |
| Pharmacist | 28(3.7) |
| Other<sup>(*)</sup> | 10(1.3) |
| Year in practice |  |
| < 5 years | 180 (23.6) |
| 5–10 years | 214(28.0) |
| 11–20 years | 223 (29.3) |
| > 20 years | 146 (19.1) |
| Practice setting |  |
| Hospital-based (government) | 325 (42.6) |
| Hospital-based (private) | 358(46.9) |
| Private practice | 160(21.0) |
| Academic/research | 91(11.9) |
| Country |  |
| Vietnam | 270 (35.4) |
| Philippines | 172 (22.6) |
| **Singapore** | 23 (2.9) |
| Thailand | 130 (17.0) |
| Malaysia | 105 (13.8) |
| Indonesia | 63 (8.3) |
| Proportion of prescriptions containing PPIs in the past month |  |
| ≤ 10% | 55(7.2) |
| ≥ 51% | 170 (22.3) |
| 11–20% | 102 (13.4) |
| 21–30% | 169 (22.1) |
| 31–40% | 143 (18.7) |
| 41-50% | 124 (16.3) |

(\*) *Included: Occupational Medicine (3), Dermatology (2), Emergency Medicine (2), Ophthalmology (1), Pediatrics (1), Traditional Medicine (1)*

The most frequently reported indications for long-term PPI use were gastroesophageal reflux disease (GERD) (67.8%), refractory GERD (57.0%), and antiplatelet therapy-induced gastric protection (50.7%) (Table 2). GIs were more likely than non-GIs to prescribe PPIs for Barrett’s esophagus (58.9% vs 29.3%, *p*<0.001), refractory GERD (68.4% vs 45.9%, *p*<0.001), antiplatelet therapy-induced gastric protection (62.3% vs 37.7%, *p*<0.001), and nonsteroidal anti-inflammatory drug (NSAID)-induced gastric protection (38.2% vs 29.3%, *p*=0.01). Interestingly, HEs were more likely than their LE counterparts to prescribe PPIs for chronic steroid therapy-induced gastric protection (35.2% vs 26.4%, *p* = 0.008).

**Table 2.**
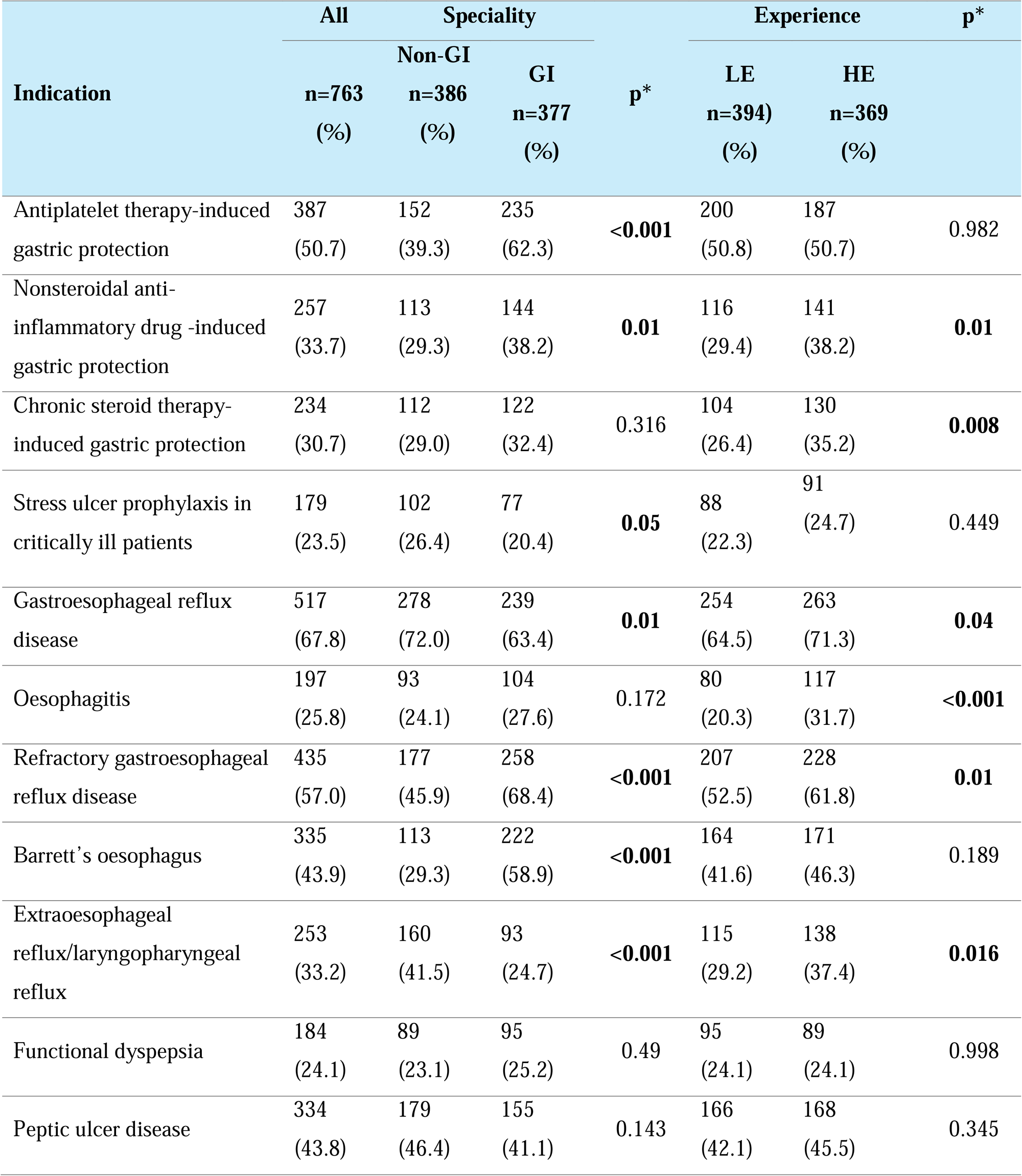
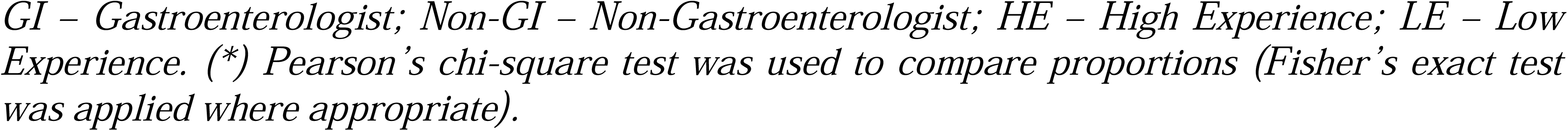
Common indications for long-term proton pump inhibitor use.

Regarding patterns of PPI prescribing, majority of respondents used standard, guideline-recommended dosages (70.1%) and usually for short-term (68.9%) (Table 3). GIs were more likely than non-GIs to prescribe combination therapy (70.8% vs 49.0%, *p* < 0.001), to titrate dosages based on symptoms (56.5% vs 44.0%, *p* < 0.001), and to use PPIs long-term (54.9% vs 43.0%, *p* < 0.001). Similarly, compared to LE providers, HE providers used combination therapy (65.9% vs 54.1%, *p*<0.001), symptom-based titration (55.3% vs 45.4%, *p* = 0.008), and long-term treatment (53.4% vs 44.7%, *p* = 0.016) more frequently.

**Table 3.**
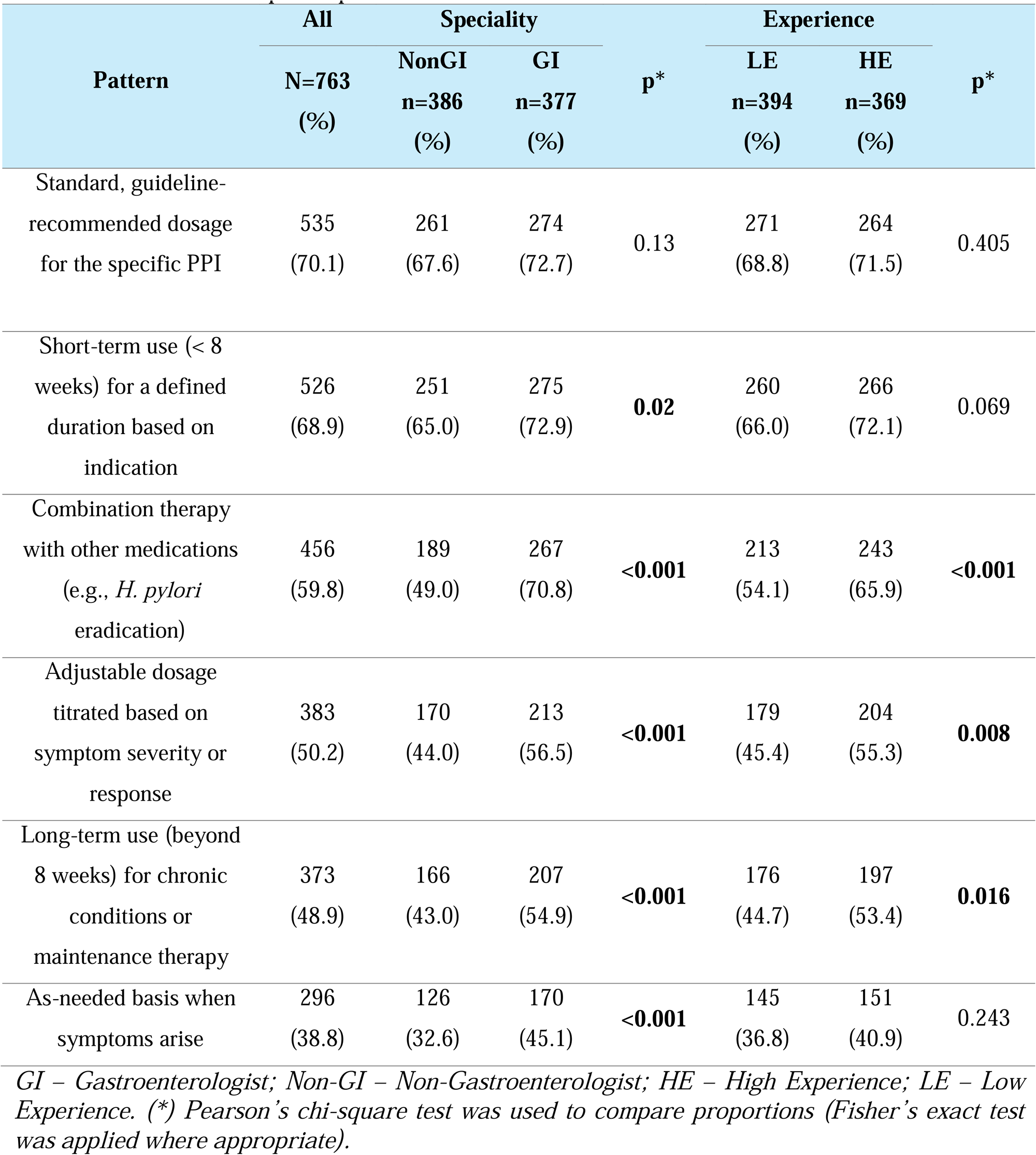
Patterns of PPI prescription or recommendation.

The most common concerns for long-term PPI use were cost effectiveness (67.8%), side effects (53.9%), and potential drug interactions (51.2%). GIs expressed significantly greater concern on side effects than the non-GIs (60.7% vs 47.2%, *p*<0.001). HE providers reported more concerns about side effects (60.7% vs 47.5%, *p*<0.001) and patient compliance (55.0% vs 42.6%, *p*<0.001) than their LE counterparts (Table 4). More than half of participants (59.2%) reported reassessing the indication for PPI use at every follow-up visit. The patterns of reassessment did not differ significantly either between GIs and non-GIs or between HE and LE healthcare providers (Table 5).

**Table 4.**
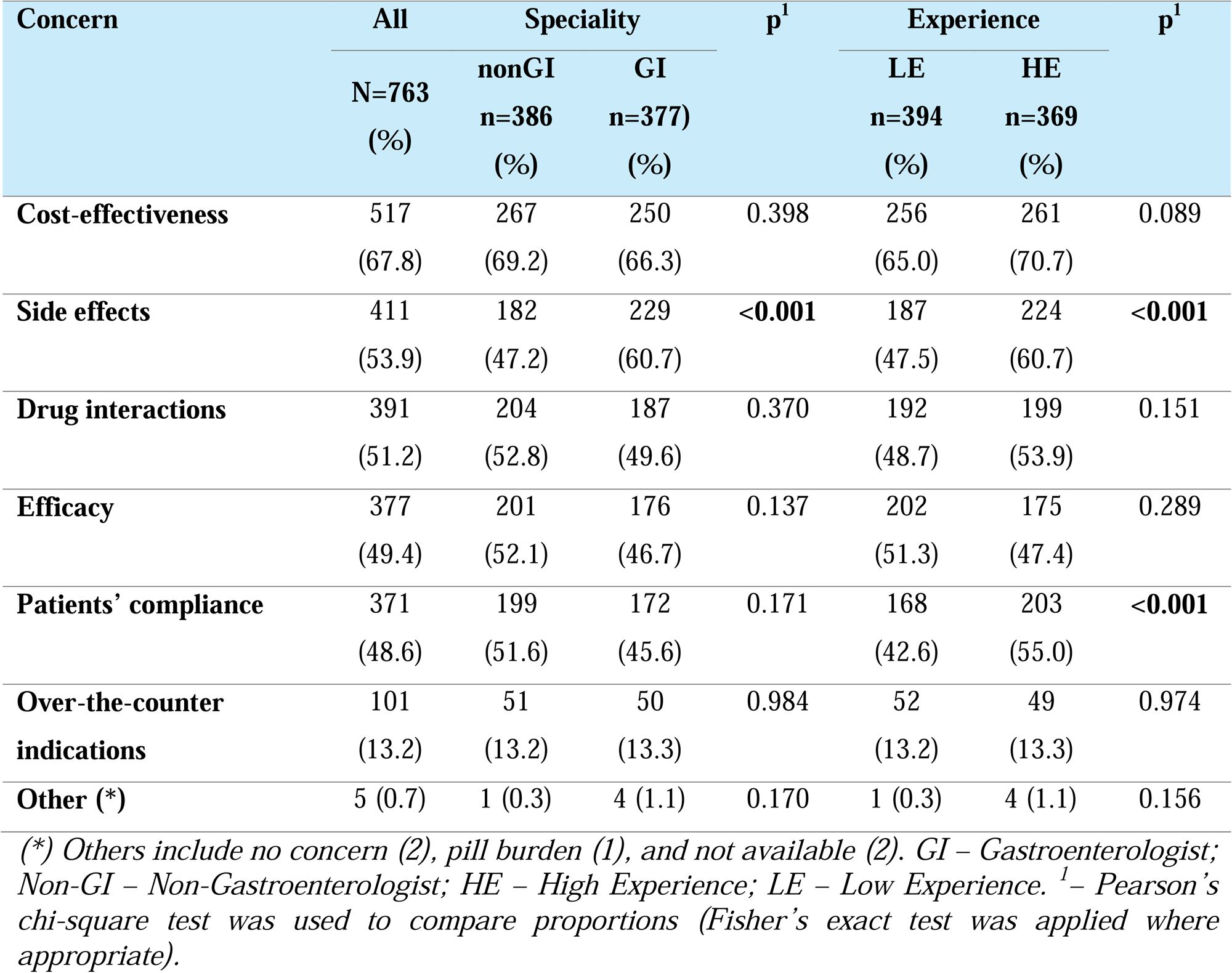
Concerns of participants when prescribing or recommending long-term PPI therapy.

| Concern | All | Speciality |  | p<sup>1</sup> | Experience |  | p<sup>1</sup> |
| --- | --- | --- | --- | --- | --- | --- | --- |
|  | N=763<br>(%) | nonGI<br>n=386<br>(%) | GI<br>n=377<br>(%) |  | LE<br>n=394<br>(%) | HE<br>n=369<br>(%) |  |
| **Cost-effectiveness** | 517<br>(67.8) | 267<br>(69.2) | 250<br>(66.3) | 0.398 | 256<br>(65.0) | 261<br>(70.7) | 0.089 |
| **Side effects** | 411<br>(53.9) | 182<br>(47.2) | 229<br>(60.7) | **<0.001** | 187<br>(47.5) | 224<br>(60.7) | **<0.001** |
| **Drug interactions** | 391<br>(51.2) | 204<br>(52.8) | 187<br>(49.6) | 0.370 | 192<br>(48.7) | 199<br>(53.9) | 0.151 |
| **Efficacy** | 377<br>(49.4) | 201<br>(52.1) | 176<br>(46.7) | 0.137 | 202<br>(51.3) | 175<br>(47.4) | 0.289 |
| **Patients' compliance** | 371<br>(48.6) | 199<br>(51.6) | 172<br>(45.6) | 0.171 | 168<br>(42.6) | 203<br>(55.0) | **<0.001** |
| **Over-the-counter indications** | 101<br>(13.2) | 51<br>(13.2) | 50<br>(13.3) | 0.984 | 52<br>(13.2) | 49<br>(13.3) | 0.974 |
| **Other (\*)** | 5 (0.7) | 1 (0.3) | 4 (1.1) | 0.170 | 1 (0.3) | 4 (1.1) | 0.156 |

(\*) Others include no concern (2), pill burden (1), and not available (2). GI – Gastroenterologist; Non-GI – Non-Gastroenterologist; HE – High Experience; LE – Low Experience. <sup>1</sup>– Pearson's chi-square test was used to compare proportions (Fisher's exact test was applied where appropriate).

The most common strategies for deprescribing PPIs included gradual dose reduction (46.8%), switching to on-demand use (45.9%), and stepping down to antacids/alginates (44.8%) (Table 6). Compared with non-GIs, GIs were more likely to use gradual tapering (52.5% vs 41.2%, *p*=0.002) and on-demand regimens (55.7% vs 36.3%, *p*<0.001). HE providers were more likely to step down to H2-receptor antagonists than their LE counterparts (17.3% vs 12.2%, *p*=0.044).

### Cluster analysis of PPI prescribing patterns

The optimal number of clusters was determined to be two based on the elbow plot (Figure 2A). Heatmap visualisation revealed consistent and statistically significant differences in prescribing behaviors between the two clusters (Figure 2B). The two identified clusters exhibited significant differences in guideline adherence, dosing adjustment strategies, and rationalisation practices. Accordingly, they were categorised as ‘proactive prescribers’ and ‘conservative prescribers’. (Supplement 2).

**Figure 2.**
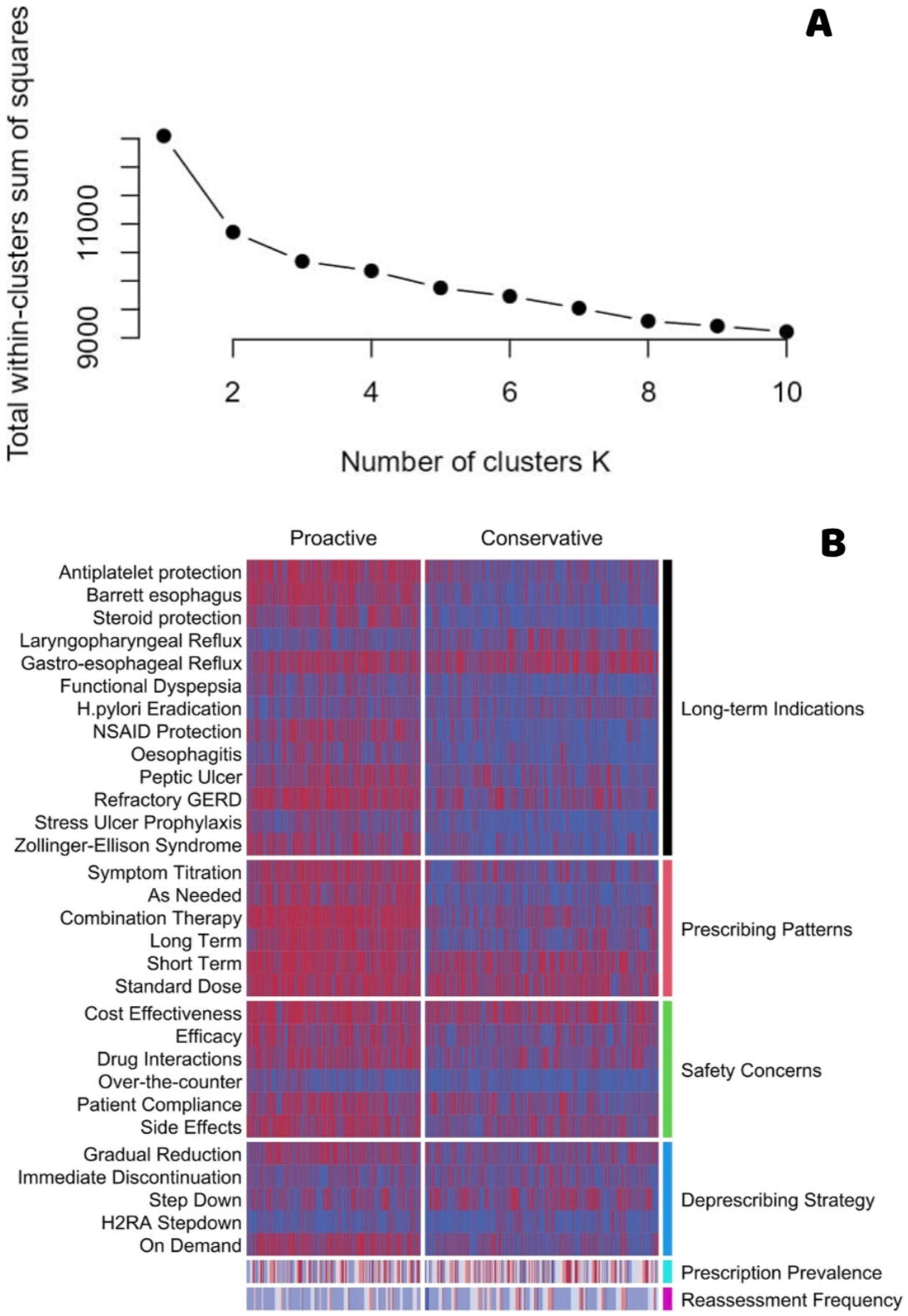
Clustering of PPI prescribing patterns. (A) The elbow plot indicates that the optimal number of clusters was 2 on the basis of within-cluster dissimilarity via the K-modes algorithm. (B) Heatmap visualisation reveals two distinct prescribing patterns: a “Proactive” cluster characterised by frequent indication-based prescribing, dose adjustment strategies, and diverse rationalisation methods and a “Conservative” cluster characterised by limited intervention and a narrower range of prescribing behaviors. Each column represents an individual respondent; rows reflect survey items on indications, adjustment, concerns, and deprescribing strategies. *Long-term Indication: Antiplatelet protection – Antiplatelet therapy-induced gastric protection; Barrett’s esophagus – Barrett’s esophagus; Steroid protection – Chronic steroid therapy-induced gastric protection; Laryngopharyngeal Reflux – Extraesophageal reflux/laryngopharyngeal reflux; Gastroesophageal Reflux – Gastroesophageal reflux disease; Functional Dyspepsia – Functional dyspepsia; H Pylori Eradication – Helicobacter pylori eradication therapy; NSAID protection – NSAID-induced gastric protection; Oesophagitis – Oesophagitis; Peptic Ulcer – Peptic ulcer disease; Refractory GERD – Refractory gastroesophageal reflux disease; Stress Ulcer Prophylaxis – Stress ulcer prophylaxis; Zollinger-Ellison syndrome – Zollinger–Ellison syndrome; Deprescribing Strategy; Symptom Titration – Symptom-based titration; As Needed – As-needed basis; Combination Therapy – Combination with other medications; Long Term – Use beyond 8 weeks; Short Term – Short-term use (<8 weeks); Standard Dose – Guideline-recommended dose; Long-term Prescribing Concerns; Cost Effectiveness – Cost-effectiveness; Efficacy – Efficacy*

Proactive prescribers (n=326, 42.7%) were characterised by a broader application of PPIs in alignment with clinical guidelines, encompassing indications such as Barrett’s esophagus, NSAID-induced gastric protection, peptic ulcer disease, and refractory GERD. They tend to employ flexible, individualised dosing approaches (e.g., symptom-based titration, combination therapy) and routinely reassess the need for ongoing therapy at each follow-up. Additionally, this group demonstrates greater familiarity with evidence-based recommendations, more frequent application of rationalisation strategies (e.g., gradual tapering, on-demand use), and increased use of adjunctive therapies such as alginates tailored to the clinical context and patient preference.

In contrast, conservative prescribers (n=437, 57.3%) used PPIs more restrictively, with fewer recognised indications and lower adherence to individualized or long-term prescribing approaches. They were less likely to reassess therapy regularly and reported limited guideline-based rationalisation strategies. Familiarity with adjunctive options—such as alginate therapy for GERD management—was also notably reduced in this group. Multivariate analysis revealed several independent predictors of proactive prescribing, including medical specialty, years of clinical experience, work setting, and country (Table 7).

## DISCUSSION

This study provides important insights into PPI prescribing and rationalisation behaviors across the SEA region and across diverse groups of healthcare professionals including GIs, geriatric physicians and pharmacists. Using a newly developed online survey with good content validity and internal consistency, our findings revealed that while a substantial proportion of healthcare professionals adopt guideline-concordant strategies, notable heterogeneity persists in terms of reassessment practices and rationalisation approaches. In addition, using K-mode clustering, the study identified the presence of two distinct prescriber profiles among the participants i.e. the proactive vs. conservative prescribers.

Regarding typical prescription of PPIs in the SEA, it is encouraging to note that majority of participants reported using standard, guideline-recommended dosages and limiting therapy to short-term duration based on the clinical indications (70.1% and 68.9%, respectively). However, there is a notable difference in the use of long-term PPIs depending on the specialty of the survey participants: GIs are more likely than non-GIs to prescribe long-term PPIs for conditions supported by guideline-based indications, such as Barrett’s oesophagus and antiplatelet-induced and NSAID-induced gastric protection. They were also significantly more likely than non-GIs to apply symptom-based titration and on-demand use. This nuanced approach may reflect the GIs’ greater familiarity with PPI pharmacodynamics and more comfortable in tailoring therapy based on individual response, as supported by previous studies ^10–12^. However, regarding long-term use of PPIs, our study showed that there were potentially inappropriate indications, especially for managing GERD (67.8%), peptic ulcer disease (43.8%) and preventing steroid-induced gastrointestinal injury (30.7%). GERD is typically mild and rarely presents with complications in Asia.^7,13^ Therefore, the latest SEA guideline recommend on-demand PPI therapy for nonerosive or mild erosive reflux disease, as it is as effective as continuous therapy.^11,13,14^ Similarly, peptic ulcer healing generally requires only 4–8 weeks of PPI treatment unless *H. pylori* eradication is unsuccessful or when aspirin/NSAID use must be continued.^11,15–17^ Although chronic steroid use has historically prompted gastroprotection, current guidelines worldwide suggest that PPI prophylaxis should be reserved for patients with increased gastrointestinal risk who are on antiplatelet or NSAID therapy, and not steroids alone.^11,12,16^ Interestingly, this practice of PPI prophylaxis was more commonly reported among HE prescribers which may reflect delayed adoption of evolving recommendations among HE prescribers, highlighting the importance of targeted continuing medical education.

Reassessment of PPI therapy is an essential practice to ensure continued clinical justification and to prevent inappropriate long-term use. In our study, most healthcare professionals (76%) reported reassessing PPI indications either at every follow-up visit or at regular intervals of 1–3 months. This frequency is consistent with international guidelines, particularly when PPI use extends beyond the initial treatment phase.^10,11,16^ Our findings also revealed a wide range of approaches to PPI deprescribing, with most clinicians preferring structured, stepwise strategies over abrupt discontinuation. The latest AGA guidelines suggest that either dose tapering or abrupt discontinuation of PPIs be considered when deprescribing PPIs, given that patients should be advised to be mindful of developing recurrent upper gastrointestinal symptoms as a consequence of rebound acid hypersecretion.^10^ In this context, alginate therapy may serve as an effective rescue option to manage breakthrough symptoms. Mechanistically, alginates form a viscous “raft” that floats on top of the gastric contents, acting as a physical barrier to prevent gastroesophageal reflux without inhibiting acid secretion—making them particularly suitable during PPI tapering. ^18,19^ Our findings reflect a generally high level of awareness of PPI stewardship across clinical domains. Nonetheless, the remaining surveyed healthcare professionals admitted to less frequent reassessment, raising concerns about missed opportunities for deprescribing.

From the K-mode cluster analysis, more than half of the survey participants were classified as conservative PPI prescribers underscored the importance and urgency of developing a regional practice guideline tailored to the specific needs and epidemiological context of the SEA region. Specifically, there were statistically significant differences in the proportion of conservative prescribers across countries. With respect to medical specialties, GIs were more likely to adopt proactive approaches than the non-GIs, and HE providers were more frequently proactive than their LE counterparts. A recent study conducted in Malaysia investigating factors influencing healthcare providers’ behaviours in deprescribing revealed that their knowledge and clinical experience played pivotal roles in shaping decisions to reduce or discontinue medications.^20^ However, this study addressed general attitudes toward deprescribing without focusing specifically on PPIs. Our findings corroborate these results within the specific context of PPI use. A recent systematic review highlighted various strategies can effectively support PPI deprescription, including interprofessional collaboration, the application of clinical decision-making algorithms, and active patient involvement^21^; hence, our findings contribute to identifying target subgroups of healthcare providers that should be prioritized to improve the appropriate prescribing and deprescribing of PPIs.

An important strength of our study lies in its focus on healthcare professionals’ perspectives regarding PPI use and rationalisation, which is based on a relatively large and diverse sample. This approach provides a broader understanding than prior studies in the region, which have primarily relied on prescription audits. Additionally, the validated survey and clustering analysis yield robust insights into prescriber behaviour, offering context-specific guidance for policy and clinical practice. However, this study has several limitations. First, its generalizability is constrained by the fact that participants were drawn from only six of the ten countries in the SEA region. Second, the unequal distribution of respondents across countries may introduce bias and limit the representativeness of PPI prescribing and rationalisation practices across the region. These limitations should be considered when interpreting and applying the findings to regional policy development. Third, this study relied on self-reported survey responses, which may be subjected to recall or social-desirability bias. To address this, future studies could incorporate objective data, such as prescription records, to validate findings.

In conclusion, this study revealed marked variability in PPI prescribing and rationalisation practices across the SEA region. It also identified several indications where PPI use are likely inappropriate, both in terms of clinical justification and prolonged duration of therapy. Most importantly, the study characterized a distinct group of conservative prescribers, who accounted for more than half of the survey respondents. These findings underscore the urgent need to establish a regional consensus to promote evidence-based clinical practice. The study may also provide an important foundation for designing targeted, context-specific interventions aimed at minimising the inappropriate and unnecessarily prolonged use of PPIs.

## Supporting information

Supplemental Digital Content 1

## ACKNOWLEDGEMENT

The authors extend their gratitude to all healthcare professionals across Southeast Asia who participated in the survey and contributed to this study. ChatGPT (OpenAI) was used solely to enhance the fluency and clarity of the English language in this manuscript. It was not involved in generating, analysing, or interpreting any scientific content. All elements of study design, data collection, statistical analysis, and interpretation of results were independently conducted and rigorously verified by the authors.

## Author contributions

DTQ: Conceptualization, Methodology, Validation, Data curation, Formal analysis, Investigation, Writing – original draft, Writing – review & editing. HTM: Methodology, Validation, Data curation, Formal analysis, Software, Writing – review & editing. SL, YYL: Project administration, Supervision, Investigation, Resources, Funding acquisition, Writing – review & editing. AG, HD, SC, PK, RK, AO, WC, KS, ST, JS, AS, TC: Investigation, Writing – review & editing.

## Data sharing statement

De-identified individual participant data and the study protocol will be available upon reasonable request to the corresponding author. Data access will be granted solely for research purposes and in accordance with ethical approvals and applicable data protection regulations.

