## Supplemental Digital Content 1 for "Proton Pump Inhibitors Prescribing Behaviors and Rationalisation Strategies among Healthcare Providers in the Southeast Asia"

**Supplement 1.** Survey contents

### **Section 1: General Information**

1. **Specialty:**
   - Gastroenterology and hepatology
   - General practitioner/family physician
   - Geriatric medicine
   - Internal medicine
   - Otolaryngology
   - Pharmacist
   - Other (please specify): _________
2. **Years in practice:**
   - < 5 years
   - 5–10 years
   - 11–20 years
   - > 20 years
3. **Practice setting: (please select all that apply)**
   - Private practice
   - Hospital-based (private)
   - Hospital-based (government)
   - Academic/research
   - Other (please specify): _________
4. **Country of practice:**
   - Indonesia
   - Malaysia
   - Philippines
   - Singapore
   - Thailand
   - Vietnam
   - Other (please specify): ________

### **Section 2: PPIs Prescribing Habits**

5. **What are your common indications for prescribing or recommending PPIs long-term?**
   **(Please select all that apply)**

□	Antiplatelet therapy-induced gastric protection

□	Barrett’s oesophagus

□	Chronic steroid therapy-induced gastric protection

□	Extraesophageal reflux/laryngopharyngeal reflux (LPR)

□	Gastroesophageal reflux disease (GERD)

□	Functional dyspepsia

□	*Helicobacter pylori* eradication therapy

□	Nonsteroidal anti-inflammatory drug (NSAID)-induced gastric protection

□	Oesophagitis

□	Peptic ulcer disease

□	Refractory gastroesophageal reflux disease (GERD)

□	Stress ulcer prophylaxis in critically ill patients

□	Zollinger–Ellison syndrome

□	Other (please specify): _________

6. **How do you typically prescribe or recommend PPIs for your patients?** **(Please select all that apply)**

□	Adjustable dosage titrated on the basis of symptom severity or response

□	As-needed basis when symptoms arise

□	Combination therapy with other medications (e.g., *H. pylori* eradication)

□	Long-term use (beyond 8 weeks) for chronic conditions or maintenance therapy

□	Short-term use (< 8 weeks) for a defined duration on the basis of indication

□	Standard, guideline-recommended dosage for a specific PPI

□	Other (please specify): _________

7. **What are your concerns when prescribing or recommending PPIs for your patients long-term? (Please select all that apply)**

□	Cost-effectiveness

□	Drug interactions

□	Efficacy

□	Over-the-counter indications

□	Patient compliance

□	Side effects

□	Other (please specify): _________

8. **What percentage of your prescriptions in the past month included PPIs?**

- ≤ 10%
- 11–20%
- 21–30%
- 31–40%
- ≥ 50%

### **Section 3: PPI rationalisation strategies**

9. **How frequently do you reassess the indication for PPI use in patients currently taking the medication?**
   - Annually
   - At every follow-up visit
   - Every 1–3 months
   - Every 3–6 months
   - Only when new symptoms or concerns arise
   - Rarely or never
   - Other (please specify): _________
10. **What is your typical strategy for rationalising PPIs when they are no longer needed or indicated? (Please select all that apply)**

- Gradual dose reduction over time
- Immediate discontinuation of PPIs
- Step down to antacids or alginates
- Step-down to H2 receptor antagonists (H2RAs)
- Switching to on-demand PPI use
- Other (please specify): _________

11. **How often do you consider using alginate as part of your PPI rationalisation strategy for patients with GERD in your real-life practice?**

- Always
- Often
- Occasionally
- Rarely
- Never

12. **What are the key factors to consider when deciding to use alginate as part of a PPI rationalisation strategy in GERD management? (Please select all that apply)**

□	Availability and cost of alginate therapy

□	Evidence supporting its efficacy in mild-to-moderate GERD

□	Patient preference for nonsystemic/nonpharmaceutical drugs

□	PPI tolerance or contraindications

□	Recommendations from clinical guidelines

□	Risk of rebound acid hypersecretion during PPI tapering

□	Other (please specify): _______________

13. **What are the reasons for not using alginate as part of a PPI rationalisation strategy in GERD management? (Please select all that apply)**

□	Accessibility issues

□	Concerns about patient adherence to alginate therapy

□	Cost

□	Lack of evidence supporting its efficacy

□	Limited familiarity with alginate as a treatment option

□	Perceived ineffectiveness compared with other therapies

□	Preference for other treatment options (e.g., H2RA)

□	None of the above difficulties encountered

□	Other (please specify): _________

---o0o---

**Supplement 2.**

**Supplement 2A.** Comparative characteristics of proactive and conservative PPI prescribing patterns

| **Characteristic** | **Group 1 N = 326***^1^*  *^(^*Proactive Prescribers)  *n* (%) | **Group 2 N = 437***^1^*  *^(^*Conservative Prescribers)  *n* (%) | **p value*^2^*** |
| --- | --- | --- | --- |
| **What are your common indications for prescribing or recommending PPIs long-term?** | | | |
| Antiplatelet therapy-induced gastric protection | 247 (75.8) | 140 (32.0) | <0.001 |
| Nonsteroidal anti-inflammatory drug-induced gastric protection | 196 (60.1) | 61 (14.0) | <0.001 |
| Chronic steroid therapy-induced gastric protection | 181 (55.5) | 53 (12.1) | <0.001 |
| Stress ulcer prophylaxis in critically ill patients | 129 (39.6) | 50 (11.4) | <0.001 |
| Gastroesophageal reflux disease (GERD) | 226 (69.3) | 291 (66.6) | 0.424 |
| Oesophagitis | 132 (40.5) | 65 (14.9) | <0.001 |
| Refractory GERD | 253 (77.6) | 182 (41.6) | <0.001 |
| Barrett’s oesophagus | 229 (70.2) | 106 (24.3) | <0.001 |
| Extraoesophageal reflux/laryngopharyngeal reflux (LPR) | 85 (26.1) | 168 (38.4) | <0.001 |
| Functional dyspepsia | 117 (35.9) | 67 (15.3) | <0.001 |
| Peptic ulcer disease | 198 (60.7) | 136 (31.1) | <0.001 |
| *Helicobacter pylori* eradication therapy | 101 (31.0) | 119 (27.2) | 0.258 |
| Zollinger-Ellison Syndrome | 192 (58.9) | 82 (18.8) | <0.001 |
| **How do you typically prescribe or recommend PPIs for your patients?** | | | |
| Standard, guideline-recommended dosage for the specific PPI | 264 (81.0) | 271 (62.0) | <0.001 |
| Short-term use (< 8 weeks) for a defined duration based on indication | 263 (80.7) | 263 (60.2) | <0.001 |
| Combination therapy with other medications (e.g., *H. pylori* eradication) | 289 (88.7) | 167 (38.2) | <0.001 |
| Adjustable dosage titrated based on symptom severity or response | 237 (72.7) | 146 (33.4) | <0.001 |
| Long-term use (beyond 8 weeks) for chronic conditions or maintenance therapy | 244 (74.8) | 129 (29.5) | <0.001 |
| As-needed basis when symptoms arise | 216 (66.3) | 80 (18.3) | <0.001 |
| **What are your concerns when prescribing or recommending PPI for your patients long-term?** | | | |
| Cost-effectiveness | 246 (75.5) | 271 (62.0) | <0.001 |
| Side effects | 226 (69.3) | 185 (42.3) | <0.001 |
| Efficacy | 219 (67.2) | 172 (39.4) | <0.001 |
| Drug interactions | 213 (65.3) | 164 (37.5) | <0.001 |
| Patients’ compliance | 203 (62.3) | 168 (38.4) | <0.001 |
| Over-the-counter indications | 66 (20.2) | 35 (8.0) | <0.001 |
| **What percentage of your prescriptions in the past month included PPIs?** | | | |
| ≤ 10% | 18 (5.5) | 37 (8.5) | <0.001 |
| ≥ 51% | 105 (32.2) | 65 (14.9) |  |
| 11–20% | 25 (7.7) | 77 (17.6) |  |
| 21–30% | 60 (18.4) | 109 (24.9) |  |
| 31–40% | 61 (18.7) | 82 (18.8) |  |
| 41-50% | 57 (17.5) | 67 (15.3) |  |
| **What is your typical strategy for rationalising PPIs when they are no longer needed or indicated?** | | | |
| Gradual dose reduction over time | 192 (58.9) | 165 (37.8) | <0.001 |
| Immediate discontinuation of PPI | 105 (32.2) | 131 (30.0) | 0.509 |
| Step down to antacids or alginates | 136 (41.7) | 206 (47.1) | 0.136 |
| Step down to H2 receptor antagonists (H2RA) | 52 (15.9) | 60 (13.7) | 0.391 |
| Switch to on-demand PPI use | 219 (67.2) | 131(30.0) | <0.001 |
| **How frequent do you reassess the indication for PPI use in patients currently taking the medication?** | | | |
| Annually | 5 (1.5) | 15 (3.4) | <0.001 |
| At every follow-up visit | 221 (67.8) | 231 (52.9) |  |
| Every 1-3 months | 47 (14.4) | 81 (18.5) |  |
| Every 3-6 months | 27 (8.3) | 43 (9.8) |  |
| Only when new symptoms or concerns arise | 25 (7.7) | 57 (13.0) |  |
| Rarely or never | 1 (0.3) | 10 (2.3) |  |
| *^1^* n (%) | | | |
| *^2^* Pearson’s Chi-squared test | | | |

**Supplement 2B.** Distribution of prescribing patterns by demographic and professional characteristics

| **Characteristic** | **All** | **Group 1 N = 326**  *^(^*Proactive Prescribers)  *n* (%) | **Group 2 N = 437**  Conservative Prescribers)  *n* (%) | **p value***^2^* |
| --- | --- | --- | --- | --- |
| **Country of practice** | | | | |
| Vietnam | 270 | 75 (23.0) | 195 (44.6) | <0.001 |
| Indonesia | 63 | 34 (10.4) | 29 (6.6) |  |
| Malaysia | 105 | 55 (16.9) | 50 (11.8) |  |
| Philippines | 172 | 68 (20.9) | 104 (23.8) |  |
| Singapore | 23 | 16 (4.9) | 7 (1.6) |  |
| Thailand | 130 | 78 (23.9) | 52 (11.9) |  |
| **Specialty** | | | | |
| Non-Gastroenterologist | 396 | 123 (37.7) | 263 (60.2) | <0.001 |
| Gastroenterologist | 377 | 203 (62.3) | 174 (39.8) |  |
| **Practice experience** | | | | |
| ≤ 10 years (LE) | 394 | 146 (44.8) | 248 (56.8) | 0.001 |
| ≥ 10 years (HE) | 369 | 180 (55.2) | 189 (43.2) |  |
| **Site of practice** | | | | |
| Academic/research | 89 | 62 (19.0) | 29 (6.6) | <0.001 |
| Hospital-based (government) | 351 | 157 (48.2) | 201 (46.0) |  |
| Hospital-based (private) | 321 | 141 (43.3) | 184 (42.1) |  |
| Private practice | 153 | 82 (25.2) | 78 (17.8) |  |

^2^ – Pearson’s chi-square test was used to compare proportions between clusters (Fisher’s exact test was applied where appropriate), LE- Low-exeprience, HE- High-experience.
